# CSF Tap Test Parameters and Short-Term Outcomes in operated and non-operated patients with idiopathic Normal Pressure Hydrocephalus: A Cohort Study

**DOI:** 10.1101/2025.08.31.25334780

**Authors:** Sagar Poudel, Deepa Dash, Aparna Wagle Shukla, Ajay Garg, Ashish Dutt Upadhyay, Naveet Wig, Roopa Rajan, Animesh Das, MR Divya, Manjari Tripathi, Achal K. Srivastava, SS Kale, P Sarat Chandra, Ashish Suri, Pramod K Pal, Hrishikesh Kumar, Sakoon Saggu, Deepti Vibha, Rajesh Kumar Singh, Jasmine Parihar, Ranveer Singh Jadon, Ved Prakash Meena, Bindu Prakash, Arunmozhimaran Elavarasi

## Abstract

**Background:** Normal Pressure hydrocephalus (NPH) is treated by ventriculoperitoneal shunting. The cerebrospinal fluid tap test (CSF-TT) is widely used to identify candidates for shunt surgery in idiopathic NPH (iNPH). This study aimed to compare the CSF tap test responses and 24-week functional outcomes between patients with probable iNPH who underwent surgery and those who did not.

**Methods:** This Ambispective cohort study included 40 patients with probable iNPH, as defined by the 2019 Japanese guidelines, from 2019 to 2024. All patients underwent a large-volume CSF-TT, and they were offered surgery based on the clinico-radiologic profile.

**Results:** Twenty-four patients underwent ventriculoperitoneal shunt surgery, and 16 did not for various reasons. No significant differences were found in the baseline or 24-hour post-CSF tap test parameters between the groups. However, at 24 weeks, 62.5% of the operated patients showed at least a 1-point improvement in mRS. In contrast, only 14.3% in the non-operated group did, indicating the beneficial role of VP shunting in NPH. Those who were operated had a 10 times higher odds (95% CI 1.6-105) of achieving at least one point improvement in mRS at 24 weeks. Interestingly, 46.7% of patients whose mRS did not improve after CSF-TT but who underwent surgery still benefited from shunting.

**Conclusion:** Shunt surgery leads to a favorable short-term functional outcome in patients with probable iNPH. However, the CSF-TT alone lacks sufficient discriminatory power to guide surgical decisions in a cohort of patients with clinico-radiological probable iNPH. A negative tap test should not preclude surgery in patients with supportive clinical and imaging features.

## Introduction

Normal pressure hydrocephalus (NPH) is a disorder characterized by higher-order gait dysfunction, cognitive dysfunction, and urinary dysfunction. It is a treatable condition with a ventriculoperitoneal (VP) shunt being the treatment of choice.^1^ The CSF tap test (CSF-TT) has been considered an important ancillary test to determine a patient’s candidacy for shunt surgery.^2,3^ The decision to subject a patient to the CSF-TT is based on clinico-radiologic features such as the triad of gait, cognitive, and urinary symptoms. The SINPHONI2 trial demonstrated that in patients with probable idiopathic NPH (iNPH), as defined by the Japanese guidelines, delaying lumbo-peritoneal shunt surgery, regardless of the CSF-TT results, led to poorer outcomes.^1,4^ A nationwide survey conducted among Indian Neurologists showed marked heterogeneity, with several respondents referring patients for surgery only if the CSF-TT was positive.^5^ But 95% of respondents reported performing the tap test, and most series have performed the CSF-TT in all patients, irrespective of whether it is used for decision-making. In this paper, we report the short-term outcomes of ventriculoperitoneal shunting in a cohort of patients with probable iNPH who underwent shunting, irrespective of CSF-TT results, and compare them with those who were considered for surgery but did not undergo shunting for various reasons. We also compared the response to the CSF-TT between these groups.

## Methods

This was an ambispective cohort study in which patients presenting to a tertiary care center in Northern India from 2019 to 2024 with possible NPH according to the Japanese guidelines^1^, with at least one of: gait disturbance, cognitive or urinary disturbances, were screened. The study was approved by the Institutional Review Board. The study was designed and reported in accordance with the STROBE reporting guidelines.

Demographics, clinical features, medical treatment, baseline gait, cognitive and urinary parameters were recorded in a structured case record form. Patients underwent baseline and 24-hour post-CSF-TT (30-50 mL of CSF drained) evaluations of gait, urinary, and cognitive function using the Boon’s gait scale, modified Rankin scale(mRS), timed up and go (TUG) test, Montreal Cognitive Assessment (MoCA), and the iNPH scale, as per the Institute’s protocol. The methodology and pre- and post-CSF-TT assessment have been described in detail elsewhere.^6^ Patients who had probable iNPH (Box 1) as per Japanese guidelines^1^ were offered a programmable VP shunt irrespective of the results of the CSF-TT. The imaging findings, correlation of cerebrospinal fluid opening pressures with CSF-TT outcomes, and shunt responsiveness in this cohort of patients^6^ have been reported in other publications.

### Box 1: Diagnostic certainty of iNPH as per Japanese guidelines^1^per Japanese guidelines

**Possible iNPH**

Possible iNPH is diagnosed if the following criteria are met:

1. More than one symptom in the clinical triad: gait disturbance, cognitive impairment, and urinary incontinence
2. The above-mentioned clinical symptoms cannot be completely explained by other neurological or non-neurological diseases.
3. Preceding diseases possibly causing ventricular dilation (including subarachnoid hemorrhage, meningitis, head injury, congenital/developmental hydrocephalus, and aqueductal stenosis) are not obvious.

**Probable iNPH**

Probable idiopathic normal pressure hydrocephalus (iNPH) is diagnosed if a patient exhibits all of the following three features.

1. Meets the requirements for possible iNPH
2. CSF pressure of 200 mmH2O or less and normal CSF content
3. One of the following two investigational features:
  a. Neuroimaging features of narrowing of the sulci and subarachnoid space over the high-convexity/ midline surface (DESH) with gait disturbance: small stride, shuffle, instability during walking, and an increase in instability on turning
  b. Improvement of symptoms after CSF tap test and/or drainage test

**Definite iNPH**

When objective improvement is seen after shunt surgery

### Short-term outcome

The patients were followed up, and outcomes at 24 weeks after surgery were assessed using the modified Rankin scale (mRS), in clinic or telephonically, as per the patient’s preference. In patients who did not undergo surgery, outcome assessment was performed 24 weeks following the CSF-TT. In the retrospective arm, telephonic interviews were conducted to assess the patient or caregiver recall based on the mRS at 24 weeks. We defined shunt responsiveness as a reduction of at least 1 point on the mRS at 24 weeks after the VP shunt surgery. For prospectively recruited patients, we also assessed the iNPH score at 24 weeks, and a one-point reduction was considered indicative of shunt responsiveness.

### Statistical analysis

Data were collected and managed using REDCap (Research Electronic Data Capture), a secure, web-based application designed to support data capture for research studies, hosted by the All India Institute of Medical Sciences (AIIMS), New Delhi, India. Data were exported and cleaned using Microsoft Excel (Microsoft Corp., Redmond, WA, USA). Descriptive statistics, including frequencies and percentages, were used to describe categorical variables. Means and standard deviations were used for continuous variables with normal distributions, while medians and interquartile ranges were used for nonparametric data. For comparisons between groups, an independent samples t-test was used for normally distributed data, and the Mann–Whitney U test was used for non-normally distributed data. Dichotomous variables were compared using the Chi-squared test or Fisher’s exact test. Odds ratios (ORs) and 95% confidence intervals (CIs) for good short-term outcomes were computed using Fisher’s exact test, which provides exact CIs appropriate for small sample sizes. All statistical analyses were performed using Stata version 17 (StataCorp, College Station, TX, USA). A p-value of <0.05 was considered statistically significant.

## Results

We screened 48 patients with possible iNPH: 20 in the retrospective arm (2019-2022) and 28 in the prospective arm. After clinical assessment, eight patients were excluded as they were unlikely to have iNPH - two had secondary NPH, three had vascular cognitive impairment, two were diagnosed with progressive supranuclear palsy, and one had coexisting Parkinson’s disease and Alzheimer’s disease. Forty patients with probable iNPH underwent a CSF-TT, out of whom 24 (7 from the prospective arm) underwent VP shunt surgery, and 16 (13 from the prospective arm) did not for various reasons – four were awaiting surgery, three were surgically unfit, and nine refused consent. The median duration to VP shunt after CSF-TT was 22.5 days (IQR 12.5-65). There were no significant differences in demographics, clinical features, or various scores between the groups at baseline; the results are summarized in Table 1 and Supplementary Table 1.

**Table 1:** Baseline features of patients with probable iNPH who underwent the CSF-TT.

| Parameter | Operated (n=24) | Non-Operated (n=16) | p-value |
| --- | --- | --- | --- |
| Age (years), mean± SD | 64.1 ± 9.5 | 64.8 ± 8.2 | 0.79 |
| Gender (male), n(%) | 20 (83.3%) | 15 (93.8%) | 0.63 |
| Symptom duration (months), median (IQR) | 24 (18.5-42) | 36(15-48) | 0.71 |
| Gait dysfunction, n(%) | 24 (100%) | 15 (93.8%) | 0.4 |
| Cognitive dysfunction, n(%) | 22 (91.7%) | 15 (93.8%) | >0.99 |
| Urinary Disturbance, n(%) | 23 (95.8%) | 13 (81.3%) | 0.28 |
| Duration of gait dysfunction(months), median (IQR) | 24 (12-36) | 24 (12-36) | 0.93 |
| Duration of cognitive dysfunction (months), median (IQR) | 19.5 (8-24) | 24 (6-48) | 0.44 |
| Duration of urinary disturbance (months), median (IQR) | 18 (8-30) | 18 (9-36) | 0.97 |
| Duration of falls (months), median (IQR) | 7 (4-36) | 12 (6-24) | 0.63 |
| iNPH gait score, median (IQR) | 1 (1-2.5) | 1 (1-2.5) | 0.35 |
| iNPH cognition score, median (IQR) | 1 (1-2.5) | 1 (1-2.5) | 0.36 |
| iNPH urinary score, median (IQR) | 1.5 (1-3) | 1 (1-2.5) | 0.11 |
| iNPH composite Score, median (IQR) | 5 (3-7) | 3 (2.5-5.5) | 0.18 |
| MoCA Score, mean±SD | 17.4±6.1 | 15.2± 7.0 | 0.48 |
| Modified Rankin Score (mRS), median (IQR) | 3.0 (2.0–3.0) | 3.0 (2.0–4.0) | 0.48 |
| mRS 1, n(%) | 0 | 1 (6.2%) | 0.54 |
| mRS 2, n(%) | 6 (37.5%) | 6 (25%) |  |
| mRS 3 , n(%) | 10 (41.7%) | 6 (37.5%) |  |
| mRS 4, n(%) | 7 (29.2%) | 2 (12.5%) |  |
| mRS 5, n(%) | 1 (4.2%) | 1 (6.3%) |  |
| Timed Up and Go (TUG) (sec), mean± SD | 30.6±11.2 | 28.2±13.6 | 0.67 |
| Falls, n(%) | 11 (45.8%) | 4 (25%) | 0.32 |
| Levodopa responsiveness ,n(%) (n=4) | 1 (25%) | 1 (25%) | >0.99 |
| Hypertension | 15 (62.5%) | 8 (50%) | 0.43 |
| Diabetes | 15 (62.5%) | 6 (37.5%) | 0.12 |
| Stroke | 4 (16.7%) | 0 (0%) | 0.09 |
| Coronary artery disease | 2 (8.3%) | 4 (25%) | 0.15 |
| Dyslipidemia | 2 (8.3%) | 1 (6.25%) | 0.81 |
| Benign Hypertrophy of the Prostate | 2 (8.3%) | 0 (0) | 0.24 |

We also examined gait scores and components, as well as urinary and cognitive scales, at baseline and 24 hours after the CSF-TT. We found no significant difference between the operated and non-operated groups. (Supplementary Table 2) We also found no difference between the operated and non-operated groups in terms of changes in Boon’s gait score or its components, including total walk score, step score, time score, iNPH composite score, MoCA score, TUG test score, and mRS following the CSF-TT. (Table 2)

**Table 2:** Changes in gait parameters, iNPH score 24 hours after CSF tap test.

|  | **Operated (n=24)** | **Non-operated (n=16)** | **P value** |
| --- | --- | --- | --- |
| **Walking Independently before CSF-TT, n/N(%)** | 18/24 (75%) | 13/16 (81.3%) | 0.72 |
| **Improvement in tandem walking, n/N(%)** | 3/18 (16.7%) | 0 | 0.24 |
| **Improvement in turning, n/N(%)** | 9/18 (50%) | 3/13 (23.1%) | 0.16 |
| **Improved Trunk Balance, n/N(%)** | 2/18 (11.1%) | 1/13 (7.7%) | >0.99 |
| **Improvement in Wide-Based Stride, n/N(%)** | 5/18 (27.8%) | 2/13 (15.4%) | 0.67 |
| **Improvement in Small Steps, n/N(%)** | 8/18 (44.4%) | 4/13 (30.8%) | 0.48 |
| **Improved Foot Clearance, n/N(%)** | 7/18 (38.9%) | 4/13 (30.8%) | 0.72 |
| **Improved Start Hesitation, n/N(%)** | 6/18 (33.3%) | 1/13 (7.7%) | 0.19 |
| **Resolved Tendency to Fall, n/N(%)** | 2/18 (11.1%) | 2/13 (15.4%) | >0.99 |
| **Able to walk unassisted from pre-tap assisted walking/inability to walk, n/N(%)** | 2/6 (33.3%) | 1/3 (33.3%) | >0.99 |
| **Total walk score change, median (IQR)** | 2 (0-6) | 2 (0-3) | 0.19 |
| **Steps score change, median (IQR)** | 2 (0-3) | 0 (0-2) | 0.06 |
| **Time score change, median (IQR)** | 1(0-2) | 1 (0-3) | 0.79 |
| **Gait Scale Total Score change, median (IQR)** | 5 (2-11) | 3 (0-7.5) | 0.19 |
| **MoCA change, median(IQR)** | 3 (1-5) | 3.5 (1-5) | 0.89 |
| **TUG change, median(IQR)** | 8 (2-11) | 2.5 (0-5) | 0.11 |
| **iNPH Composite Score change, median (IQR)** | 1 (0-1) | 0.5 (0-1) | 0.54 |
| **Modified Rankin Score** | 0 (0-1) | 0 (0-1) | 0.86 |
| change, median (IQR) |  |  |  |

There were no intraoperative complications. Postoperatively, seven patients had complications. Aspiration pneumonia, subdural collection, and ventriculitis occurred in one patient each. In two patients, headache and vomiting occurred, which resolved with a reduction in shunt pressure. One patient had edema around the shunt and probable ventriculitis. One patient died in the postoperative setting, with no improvement in triad symptoms.

At 24 weeks follow-up (Table 3), a significant difference was observed between the operated and non-operated groups, with more patients in the operated group showing improvement in functional outcomes at 24 weeks compared to baseline. Fifteen patients showed an improvement of at least one point in mRS (seven patients with a one-point improvement and eight patients with a two-point improvement), and four patients did not worsen further until 24 weeks. However, among non-operated patients, seven (42.8%) worsened at 24 weeks, one patient died before 24 weeks, and five (35.7%) remained unchanged. Two patients in the non-operated group who had improved mRS after the CSF tap test maintained the same mRS level until the 24-week follow-up. The odds ratio for a good short-term outcome was 10.0 (95% CI 1.6 – 105; Fisher’s exact test, p = 0.006)

**Table 3:** Comparison of mRS change at 24 weeks compared to baseline.

|  | Post CSF tap test |  | P value | 24 weeks of follow-up |  | P value |
| --- | --- | --- | --- | --- | --- | --- |
|  | Operated (n=24) | Non-operated (n=16) |  | Operated (n=24) | Non-operated (n=14) |  |
| **≥1-point worsening** | 0 | 0 | >0.99 | 2(8.3%) | 6(42.9%) | 0.02 |
| **No change in MRS** | 13 (54.2%) | 9 (56.3%) |  | 4 (16.7%) | 5 (35.7%) |  |
| **1-point improvement** | 9 (37.5%) | 6 (37.5%) |  | 7 (29.2%) | 2 (14.3%) |  |
| **2-point improvement** | 2 (8.3%) | 1 (6.3%) |  | 8 (33.3%) | 0 |  |
| **Died** | 0 | 0 |  | 3 (12.5%) | 1(7.1%) |  |

At baseline, both groups predominantly fell in the mRS 3–6 range, with no statistically significant difference between them. Following the CSF tap test, both groups showed a trend toward better functional status (mRS 1–2); however, the improvement was not statistically significant. At 24 weeks, the operated group demonstrated a significantly higher proportion of patients in the mRS 1–2 category compared to the non-operated group (62.5% vs. 21.4%), indicating a favorable shift in short-term functional outcome following shunt surgery. (Figure 1)

**Figure 1.**
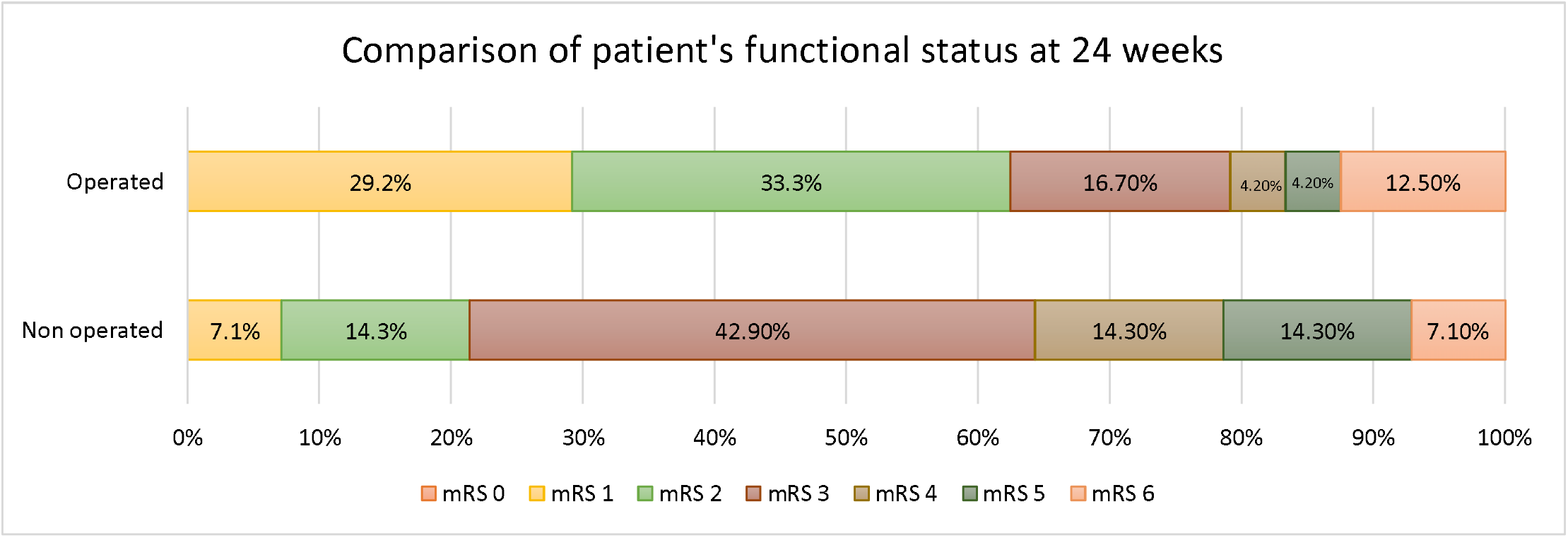
Shift analysis showing comparison of patients’ functional status at 24 weeks

## Discussion

In a highly selected cohort of patients with probable iNPH, our study found that 62.5% of patients responded to shunt surgery at 24 weeks, with no significant difference in the duration to shunt after CSF-TT between shunt responders and nonresponders. On the other hand, 16.7% were static, while 20.8% worsened despite surgery. Studies have shown that the surgical outcome ranges from 38% to 81%.^1^ In our study, a significant difference was observed in mRS change at 24 weeks between operated and non-operated patients, indicating improvement after shunt surgery. This is an interesting finding, given that there were no differences in response to large-volume CSF-TT between those who underwent surgery and those who did not. Despite the practice in various settings of referring only CSF-TT-positive patients for shunt surgery, our finding aligns with a nationwide Japanese survey, which showed a significant improvement in the modified Rankin scale among shunt-operated patients compared to the non-operated group, regardless of CSF-TT results.^7^ SINPHONI study shows 80% of shunted patients respond in terms of improvement in mRS, and 89% in terms of improvement in iNPH score.^8^ Our study revealed that surgery based on clinico-radiologic findings resulted in a response rate of 62.5% when using mRS as the outcome and 70% when using the iNPH score as the outcome. We did not find any differences in the specific CSF tap test parameters between the operated and non-operated patients. Our data did not reveal any significant differences in major parameters between the operated and non-operated patients, as assessed by Boon’s gait scale, TUG test, and iNPH scale. Only 46% of the patients who underwent surgery were CSF-TT responders, and 54% were nonresponders. And 54% of those who were not CSF-TT responders ultimately responded to the VP shunt. Had only those who responded to the CSF-TT been subjected to surgery, these patients would have missed out on the opportunity for improvement. Existing literature highlights the poor predictive value of the CSF tap test.^9^ In our recent meta-analysis, the pooled sensitivity of the CSF-TT was 67.5% (95% CI: 52-80), and the pooled specificity was 53% (95%CI: 41-66).^10^ This emphasizes that a negative tap test does not necessarily rule out shunt benefit. In our study, seven patients (46.7%) in the operated group who did not respond to the CSF tap test at 24 hours have responded to shunting. This suggests that a non-improved or borderline tap test response should not be excluded automatically from surgery if the clinical picture and imaging are strongly suggestive of iNPH. Though the existing guidelines do not recommend the exclusion of patients based on response to the CSF-TT, even the recent randomized controlled trial by Luciano et al. recruited only those who responded.^11^

Our study has several limitations. As a tertiary care institute, we often suffer from referral bias. Although the decision to refer patients for surgery was not based on the CSF-TT, it is possible that patients with probable iNPH who did not want to undergo CSF-TT or who did not consider surgery following referral were excluded from the cohort, as they did not present to our clinics for treatment. We did not use other adjunctive tests, such as the lumbar infusion test, to predict shunt responsiveness. The decision to operate was not randomized, and the reasons for no operations were variable, so selection bias is a possibility. Similarly, although some patients refused surgery due to lack of improvement, we did not specifically assess the proportion of patients with subjective lack of improvement in the operated cohort. Although we are a large-volume center, the sample size is small, which might affect the generalizability of our results. Thirdly, only functional improvement in mRS was focused on during outcome assessment. Improvement in other cognitive parameters and urinary disturbances at follow-up is unknown. We used the modified Rankin scale as an outcome measure because it is widely used, and the landmark randomized controlled study SINPHONI-2 used at least a 1-point improvement in the mRS as the primary outcome. The mRS is a global outcome assessment measure and a functional outcome scale with good interrater agreement, especially for telephonic assessment. The ambispective design might have introduced recall bias, as the 24-week follow-up relied on self-reported recall. The modified Rankin Scale is an investigator-assessed outcome based on patient or caregiver report; however, telephonic assessment relies heavily on such reports and could have introduced heterogeneity. The patients were not blinded to the intervention, which could have introduced bias in the reporting of the outcome. The outcomes assessed were functional outcomes, which may not have been sensitive to minor improvements, particularly given the presence of older individuals with multiple comorbidities and orthopedic issues, potentially leading to an underestimation of the effects of shunt surgery. Finally, the confidence intervals for the odds ratio of improvement following shunting were quite wide, reflecting imprecision in the results.

Additionally, patients’ shunt valve pressure adjustments and the timing of these adjustments were not recorded, which may have influenced the outcome assessment. Timely valve pressure programming may lead to appropriate drainage and symptom improvement. We reported the short-term functional outcomes at 24 weeks, and several studies have shown deterioration in the long term, thereby missing out the degenerative component of the disease.

## Conclusion

Our study reinforces the current available evidence that shunt surgery can be beneficial in a carefully selected subset of patients with probable idiopathic Normal Pressure Hydrocephalus (iNPH). Many patients show a good clinical response in the short term following ventriculoperitoneal (VP) shunting. However, the cerebrospinal fluid (CSF) tap test alone lacks sufficient discriminatory power to predict shunt responsiveness accurately. It may therefore be inappropriate to exclude patients from shunt surgery solely on the basis of a negative CSF tap test. Conversely, it should also be remembered that some patients with a positive CSF tap test do not go on to improve after surgery, highlighting the limitations of relying on this test alone. Moreover, currently available assessment techniques cannot reliably rule out the possibility of significant benefit after shunt placement. A comprehensive, multimodal approach—including detailed gait analysis, neuroimaging findings, and judicious use of the CSF tap test—is essential for selecting appropriate surgical candidates. Long-term outcomes following shunt surgery remain poorly understood and need to be systematically studied to inform prognosis and counsel patients.

## Data Availability

All data produced in the present study are available upon reasonable request to the authors

## Financial Disclosures for the previous 12 months

The authors declare that there are no additional disclosures to report.

## Ethical Compliance Statement

The study was reviewed and approved by the Institute Ethics Committee at the All India Institute of Medical Sciences, New Delhi (No. IECPG-222/20.4.23, RT-14/07.06.23). The participants and their legal guardians provided informed consent to participate in this study. We confirm that we have read the Journal’s position on issues involved in ethical publication and affirm that this work is consistent with those guidelines.

## References

1. Nakajima M, Yamada S, Miyajima M, et al. Guidelines for Management of Idiopathic Normal Pressure Hydrocephalus (Third Edition): Endorsed by the Japanese Society of Normal Pressure Hydrocephalus. Neurol Med Chir (Tokyo). 2021;61(2):63–97. doi:10.2176/nmc.st.2020-0292

2. Hu T, Lee Y. Idiopathic normal-pressure hydrocephalus. Can Med Assoc J. 2019;191(1):E15–E15. doi:10.1503/cmaj.180877

3. Carswell C. Idiopathic normal pressure hydrocephalus: historical context and a contemporary guide. Pract Neurol. 2023;23(1):15–22. doi:10.1136/pn-2021-003291

4. Kazui H, Miyajima M, Mori E, Ishikawa M. Lumboperitoneal shunt surgery for idiopathic normal pressure hydrocephalus (SINPHONI-2): an open-label randomised trial. Lancet Neurol. 2015;14(6):585–594. doi:10.1016/S1474-4422(15)00046-0

5. Elavarasi A, Poudel S, Dash D, et al. Current Practice Patterns Among Indian Neurologists in the Evaluation and Management of Normal Pressure Hydrocephalus: A Nationwide Cross-Sectional Survey. SSRN. Preprint posted online 2025. doi:10.2139/ssrn.5376438

6. Poudel S, Dash D, Fasano A, et al. Diagnostic Accuracy of CSF Tap-Test Parameters in Predicting Shunt Responsiveness in Normal Pressure Hydrocephalus: A Cohort Study. SSRN. Preprint posted online 2025. doi:10.2139/ssrn.5413559

7. Nakajima M, Miyajima M, Ogino I, et al. Shunt Intervention for Possible Idiopathic Normal Pressure Hydrocephalus Improves Patient Outcomes: A Nationwide Hospital-Based Survey in Japan. Front Neurol. 2018;9:421. doi:10.3389/fneur.2018.00421

8. Ishikawa M, Hashimoto M, Mori E, Kuwana N, Kazui H. The value of the cerebrospinal fluid tap test for predicting shunt effectiveness in idiopathic normal pressure hydrocephalus. Fluids Barriers CNS. 2012;9(1):1. doi:10.1186/2045-8118-9-1

9. Mihalj M, Dolić K, Kolić K, Ledenko V. CSF tap test - Obsolete or appropriate test for predicting shunt responsiveness? A systemic review. J Neurol Sci. 2016;362:78–84. doi:10.1016/j.jns.2016.01.028

10. Aliyar A, Dash D, Fasano A, et al. Accuracy of CSF Tap Test and Lumbar Infusion Test in Predicting Shunt Response in Idiopathic Normal Pressure Hydrocephalus: A Systematic Review and Meta-Analysis. medRxiv. Preprint posted online November 10, 2025:2025.11.09.25339846. doi:10.1101/2025.11.09.25339846

11. Luciano MG, Williams MA, Hamilton MG, et al. A Randomized Trial of Shunting for Idiopathic Normal-Pressure Hydrocephalus. N Engl J Med. 2025;393(22):2198–2209. doi:10.1056/NEJMoa2503109

