## Supplementary Table for "CSF Tap Test Parameters and Short-Term Outcomes in operated and non-operated patients with idiopathic Normal Pressure Hydrocephalus: A Cohort Study"

Supplementary Table 1: Baseline TUG test features between operated and non-operated patients

| Gait features | Operated (n=24) | Non operated (n=16) | P value |
| --- | --- | --- | --- |
| TUG stopped to rest,n(%) | 1 (10%) | 2 (16.7%) | >0.99 |
| Low tentative pacen(%) | 10 (41.7%) | 8 (50%) | 0.75 |
| Loss of balance,n(%) | 2 (8.3%) | 2 (12.5%) | >0.99 |
| Short strides,n(%) | 10 (61.7%) | 9 (56.3%) | 0.52 |
| Little or no arm swing,n(%) | 9 (37.5%) | 6 (37.5%) | >0.99 |
| Steadying self on walls,n(%) | 0 | 1 (6.3%) | 0.4 |
| Shuffling,n(%) | 2 (8.3%) | 1 (6.3%) | >0.99 |
| En bloc turning,n(%) | 6 (25%) | 6 (37.5%) | 0.49 |
| Not using assistive device properly,n(%) | 2 (8.3%) | 1 (6.3%) | >0.99 |

| **Parameter** | **Pre CSF tap test** | | **P value** | **Post CSF tap test** | | **P value** |
| --- | --- | --- | --- | --- | --- | --- |
|  | **Operated(n=24)** | **Non operated (n=16)** |  | **Operated** | **Non operated** |  |
| **Walking Independently, n(%)** | 18 (75%) | 13 (81.3%) | 0.72 | 20 (83.3%) | 14 (87.5%) | >0.99 |
| **Tandem Walking walking, n(%)** | 17 (94.4%) | 12 (92.3%) | >0.99 | 16 (80%) | 13 (92.9%) | 0.38 |
| **Turning Disturbed, n(%)** | 13 (72.2%) | 7 (53.9%) | 0.44 | 5 (25%) | 4 (28.6%) | >0.99 |
| **Trunk Balance disturbed, n(%)** | 8 (44.4%) | 1 (7.7%) | 0.04 | 7 (35%) | 2 (14.3%) | 0.25 |
| **Wide-Based Stride, n(%)** | 10 (55.6%) | 5 (38.5%) | 0.47 | 7 (35%) | 4 (30.8%) | >0.99 |
| **Small Steps, n(%)** | 15 (83.3%) | 8 (61.5%) | 0.22 | 10 (48.5%) | 6 (42.9%) | 0.73 |
| **Reduced Foot Clearance, n(%)** | 17 (94.4%) | 9 (69.2%) | 0.13 | 11 (55%) | 5 (35.7%) | 0.32 |
| **Start Hesitation, n(%)** | 9 (50%) | 3 (23.1%) | 0.15 | 4 (20%) | 2 (14.3%) | >0.99 |
| **Tendency to Fall, n(%)** | 6 (33.3%) | 2 (15.4%) | 0.41 | 6 (30%) | 0 | 0.03 |
| **Able to Walk assisted, n(%)** | 5 (20.8%) | 2 (12.5%) | >0.99 | 3 (12.5%) | 1 (6.3%) | >0.99 |
| **Unable to walk at all, n(%)** | 1 (4.2%) | 1 (6.3%) | >0.99 | 1 (4.2%) | 1 (6.3%) | >0.99 |
| **Total walk score, median (IQR)** | 12 (8-17) | 8 (6-10) | 0.05 | 6 (4-14) | 5 (4-9) | 0.43 |
| **Step score,** mean± SD | 7.7 ± 2.4 | 7.2 ±2.6 | 0.57 | 5.91±2.6 | 6.47±2.97 | 0.55 |
| **Time score, mean**±SD | 7.7±2.6 | 7.7±2.4 | 0.87 | 6.7±2.7 | 6.4±2.6 | 0.74 |
| **Total gait score, mean**± SD | 27.4±8.3 | 23.4±8.1 | 0.13 | 20.7±9.3 | 18.9±8.6 | 0.56 |
| **mRS1, n(%)** | 0 | 1 (6.2%) | 0.54 | 3 (12.5%) | 4 (25%) | 0.9 |
| **mRS 2** | 6 (37.5%) | 6 (25%) |  | 10 (41.7%) | 6 (37.5%) |  |
| **mRS3** | 10 (41.6%) | 6 (37.5%) |  | 6 (25%) | 4 (25%) |  |
| **mRS 4** | 7 (29.2%) | 2 (12.5%) |  | 4 (16.7%) | 2 (12.5%) |  |
| **mRS 5** | 1 (4.2%) | 1 (6.3%) |  | 1 (4.2%) | 0 |  |
| **MOCA score (n=19)** | 17.4±6.1 | 15.2 ± 7.0 | 0.48 | 20.1 ±4.5 | 18.1±7.7 | 0.52 |
| **TUG score (n=22)** | 30.6±11.2 | 28.2±13.6 | 0.67 | 23.2±6.5 | 24.7±14.1 | 0.76 |
| **iNPH gait score** | 1 (1-2.5) | 1 (1-2.5) | 0.35 | 1 (1) | 1 (0-1) | 0.31 |
| **iNPH cognition score** | 1 (1-2.5) | 1 (1-2.5) | 0.36 | 1 (0.5-2) | 1 (1) | 0.94 |
| **iNPH urinary score** | 1.5 (1-3) | 1 (1-2.5) | 0.11 | 1 (1-3) | 1 (0.5-1) | 0.05 |
| **iNPH composite score** | 5 (3-7) | 3 (2.5-5.5) | 0.18 | 4.5 (2-6) | 3 (1.5-4) | 0.16 |

Supplementary Table 2: Comparison of parameters before and after CSF tap test between operated and non-operated patients.
